# Comparison of Machine Learning Models in Predicting Mental Health Sequelae Following Concussion in Youth

**DOI:** 10.1101/2025.01.02.24319733

**Authors:** Jin Peng, Jiayuan Chen, Changchang Yin, Ping Zhang, Jingzhen Yang

**Affiliations:** Information Technology Research and Innovation, The Abigail Wexner Research Institute, Nationwide Children’s Hospital, Columbus, Ohio, USA; Computer Science and Engineering, The Ohio State University, Columbus, Ohio, USA; Department of Biomedical Informatics, The Ohio State University, Columbus, Ohio, USA; Center for Injury Research and Policy, The Abigail Wexner Research Institute, Nationwide Children’s Hospital, Columbus, Ohio, USA; Department of Pediatrics, The Ohio State University, Columbus, Ohio, USA

## Abstract

Youth who experience concussions may be at greater risk for subsequent mental health challenges, making early detection crucial for timely intervention. This study utilized Bidirectional Long Short-Term Memory (BiLSTM) networks to predict mental health outcomes following concussion in youth and compared its performance to traditional models. We also examined whether incorporating social determinants of health (SDoH) improved predictive power, given the disproportionate impact of concussions and mental health issues on disadvantaged populations. We evaluated the models using accuracy, area under the curve (AUC) of the receiver operating characteristic (ROC), and other performance metrics. Our BiLSTM model with SDoH data achieved the highest accuracy (0.883) and AUC-ROC score (0.892). Unlike traditional models, our approach provided real-time predictions at each visit within 12 months of the index concussion, aiding clinicians in making timely, visit-specific referrals for further treatment and interventions.

## Introduction

Concussion and mental health (MH) are significant public health issues that disproportionately affect youth^1-4^. Each year, approximately 2 million youths under 18 experience a concussion, a form of mild traumatic brain injury that can severely impact developing brains and negatively affect functional, cognitive, emotional, and sleep health^5-10^. Evidence suggests that concussion is likely associated with mental health conditions such as anxiety ^11-13^, depression ^14,15^, distress ^16,17^, and suicidality ^18,19^. If untreated or unmanaged, these mental health conditions can adversely affect concussion recovery and potentially lead to long-term health consequences. Therefore, early identification of youth at heightened risk for poor mental health outcomes following a concussion is crucial for effective prevention and treatment of these sequelae.

Machine learning (ML)^20,22^ has been shown to improve outcome prediction in traumatic brain injury (TBI) patients compared to classical regression approaches ^23,24^, offering advances in prediction methods ^25,26^, and accuracy ^21, 27-30^. However, existing ML models are primarily based on more severe TBIs, small sample sizes, and adult patients ^26,28,29^. To date, no study has utilized ML to derive a risk-prediction tool for mental health sequelae specifically in youth with concussion ^31-34^.

Youth from disadvantaged populations, including racial/ethnic minoritized groups, lower socioeconomic status (SES) households, rural areas, and those with limited access to care ^35-37^, are at a higher risk for both concussion and mental health conditions compared to their more socially advantaged peers ^38,39^. However, social determinants of health (SDoH) information is often missing from electronic medical records (EMRs), hindering our understanding of the predictors of mental health sequelae following concussion in youth with known health disparities. To address this gap, researchers at The Ohio State University developed the Ohio Children’s Opportunity Index (OCOI)^40^, which synthesizes over 34 variables related to neighborhood conditions and opportunities into a single index score, allowing for the assessment of a child’s SDoH at the census-tract level. Additionally, an increasing number of pediatric hospitals in the United States are screening children for Social Determinants of Health (SDoH) to capture unmet social and economic needs at a more individual level^41^. However, no prior study has leveraged both area-level and individual-level SDoH data to explore their role in predicting mental health sequelae following concussion in youth.

In this study, we employed a novel approach of Bidirectional Long Short-Term Memory (BiLSTM) networks for predicting MH sequelae following concussion in youth and compared its performance with traditional logistic regression and other machine learning models. We chose a bidirectional LSTM (BiLSTM) for our study because BiLSTMs are particularly effective at handling sequential data and tasks that require context from both the past and future. This is especially valuable in our research because: 1) some of our data are inherently sequential, such as injury and clinical symptom measures, and 2) understanding a patient’s mental health history prior to the index concussion, along with future outcomes (e.g., subsequent concussions or changes in clinical symptoms), is essential for accurately predicting mental health sequelae following the initial concussion. Given that concussion and MH problems disproportionately affect youth from disadvantaged populations, we also investigated whether incorporating both area-level and individual-level SDoH data in our models enhanced predictive power.

## Methods

### Data

This was a single-center retrospective study. This study was approved by Nationwide Children’s Hospital (NCH) Institutional Review Board (# 2030585). Data on youth ages 11 to 17 years with concussion from 1/1/2013 to 12/31/2022 were obtained from three databases: 1) Electronic medical records (EMR) at NCH, 2) Area-level SDoH data from the Ohio State University (OSU), and 3) Individual-level SDoH data collected at NCH since 2018.

### EMR at NCH

NCH uses a comprehensive and integrated set of clinical software systems developed by EPIC, a leading EMR vendor, to record and manage patient care interactively for NCH patients. Patient medical records, including demographics, encounters, orders, results, providers, care facility, and billing data, are extracted daily into EPIC’s proprietary backend Clarity Database. For the purpose of this study, we requested the following EMR data elements: patient demographics (i.e., age, gender, race), diagnosis codes, injury, and clinical characteristics, etc. The full list of EMR variables can be found in **Table 1**.

**Table 1.** Summary of Model Variables.

| Category | Number of variables | Description | Data source |
| --- | --- | --- | --- |
| Outcome (binary) | 1 | Mental health diagnosis (Yes vs. No) identified using ICD-9-CM codes: 293, 294, | Institutional EMR |

| Continued |  |  |  |
| --- | --- | --- | --- |
| Category | Number of variables | Description | Data source |
|  |  | 300, 308-313, and 316, and ICD-10-CM codes F30-F39, F40-F48, and F50-F59 |  |
| Demographics | 5 | Patient age, gender, race, language, residence rurality | Institutional EMR |
| Insurance info | 1 | Payer type (Private/Medicaid/Self-pay) | Institutional EMR |
| Pre-existing mental health problems | 1 | Mental health diagnosis prior to 1 year before the diagnosis date of the index concussion (Yes vs. No) identified using the same ICD codes listed above | Institutional EMR |
| Injury and clinical characteristics | 23 | Recorded date, total post-concussion symptom score (based on 22 individual symptoms such as headache, dizziness), neck pain, don't feel right, injury mechanism, sleep score, learning disability, attend school, pressure in head, concussion improved, anterograde amnesia, retro post trauma amnesia loss of consciousness, weakness in extremities etc. | Institutional EMR |
| Repeated concussions after the index concussion | 1 | Diagnosis date, concussion identified using ICD-9-CM codes 850.0, 850.1, 850.11, 850.12, 850.2, 850.3, 850.4, 850.5, and 850.9, and ICD-10-CM codes beginning S06.0 | Institutional EMR |
| Individual-level SDoH info | 5 | Consent to respond, needs of housing, food, transportation, utility | Institutional Patient Survey |
| Area-level SDoH info | 9 | Ohio Child Opportunity Index Score: overall, family stability, infant health, children's health, housing, access to care, education, environment, crime | Public Resource |

### Area-level SDoH Data

The OCOI 2.0^40^, developed by researchers at OSU, is a publicly available composite “opportunity” index that captures neighborhood resources influencing children’s healthy development at the census-tract level. The OCOI is based on 53 indicators across 8 domains—family stability, infant health, child health, health access, education, housing, environment, and criminal justice. These indicators were used to measure area-level SDoH (social determinants of health) and can be linked to the NCH EMR data, as all NCH patient records have been geocoded at the census-tract level and labeled with census tract IDs.

### Individual-level SDoH Data

Since 2018, NCH has implemented a SDoH screening survey of four health-related social needs (housing, food, transportation, utilities) that can impact patient health, with response categories of “YES”, “NO”, or “Declined to Answer”. Patients who responded “YES” are considered as having the need(s).

### Study Population

Our study population consisted of a retrospective cohort of patients aged 11 to 17 with a confirmed concussion diagnosis, each having at least one concussion-related medical encounter at NCH between January 1, 2013, and December 31, 2022. Concussions were defined using the International Classification of Diseases, Ninth and Tenth Revisions, Clinical Modification (ICD-9-CM and ICD-10-CM) codes: 850.0, 850.1, 850.11, 850.12, 850.2, 850.3, 850.4, 850.5, 850.9, and codes beginning with S06.0. The index concussion was determined by the date and time of the encounter. Mental health conditions were defined as having any of ICD-9-CM codes: 293, 294, 300, 308-313, 316, or ICD-10-CM codes: F30-F39, F40-F48, or F50-F59 following the index concussion. Exclusion criteria included: 1) patients assigned an ICD-9-CM or ICD-10-CM code for a more severe TBI within four weeks of the index concussion; 2) patients with significant injuries to body regions other than the brain on the same day as the index concussion; 3) patients receiving ongoing concussion treatment whose index concussion encounter occurred before January 1, 2013; and 4) patients with an ongoing mental health diagnosis or treatment within 12 months prior to the index concussion.

### Models

We modeled the prediction of mental health diagnoses at each follow-up visit within 12 months following the index concussion (**Figure 1**). For a repeated concussion or a new record of post-concussion symptoms occurring during the prediction window, a new prediction time will start for the repeated concussion or the new record of post-concussion symptoms, and the original prediction window will be censored. We first developed an interpretable deep learning model based on a bidirectional long short-term memory (BiLSTM) network^42^ (**Figure 2**). This model incorporated multi-modal data, including demographics, injury details, and encounter variables from electronic medical records (EMR), as well as SDoH data from public resources and hospital patient surveys, embedding these inputs into a unified representation space. A time embedding accounted for irregular intervals between medical encounters. The model utilized a bi-directional LSTM (Bi-LSTM) to process sequential data and generate output vectors, which were combined using global max pooling to create a patient representation vector. Finally, a fully connected layer and sigmoid function predicted the probability of mental health sequelae at each post-concussion encounter, continuously assessing risk up to 12 months post-injury. For comparison, we also developed several commonly used ML models, including Logistic Regression (LR), Multilayer Perceptron (MLP) ^43^, and recurrent neural networks (RNN) ^44^. **Table 1** summarizes all variables used in our models.

**Figure 1.**
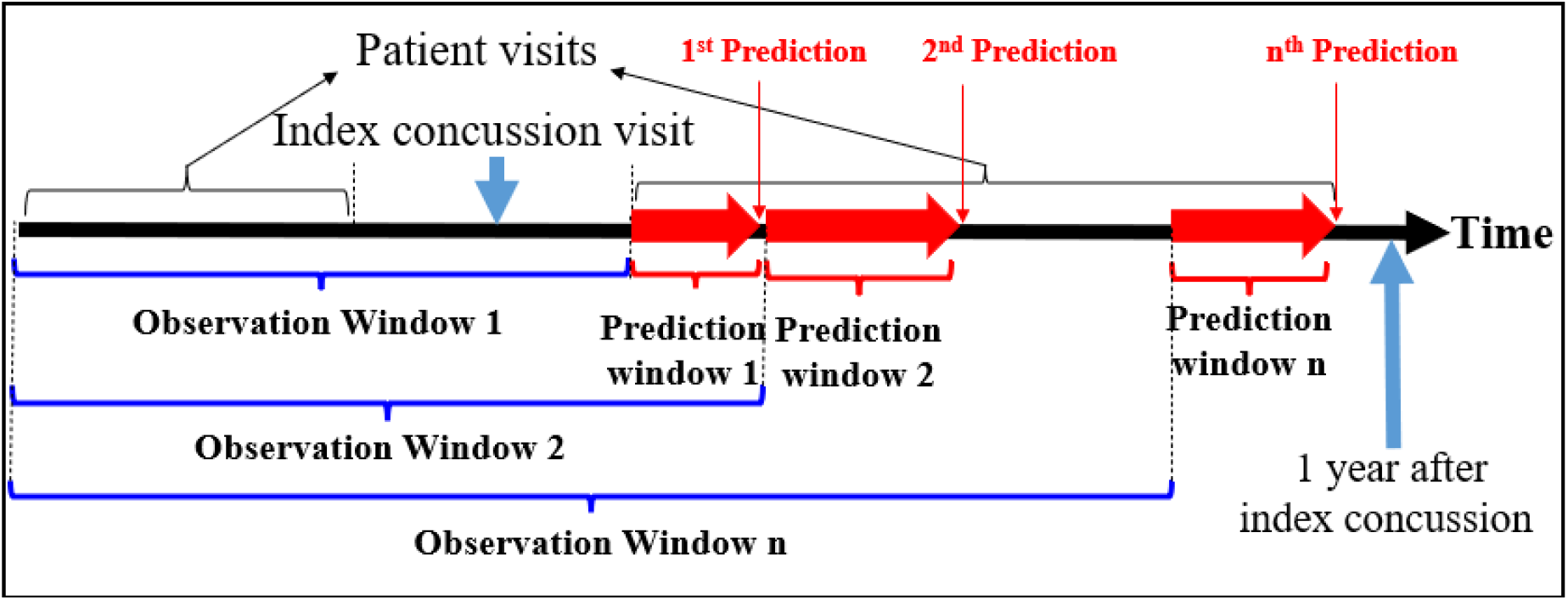
Temporal Predictions of Mental Health Diagnoses in the Year Post-Concussion. Predictions are made at each follow-up visit up to 12 months following the index concussion.

**Figure 2.**
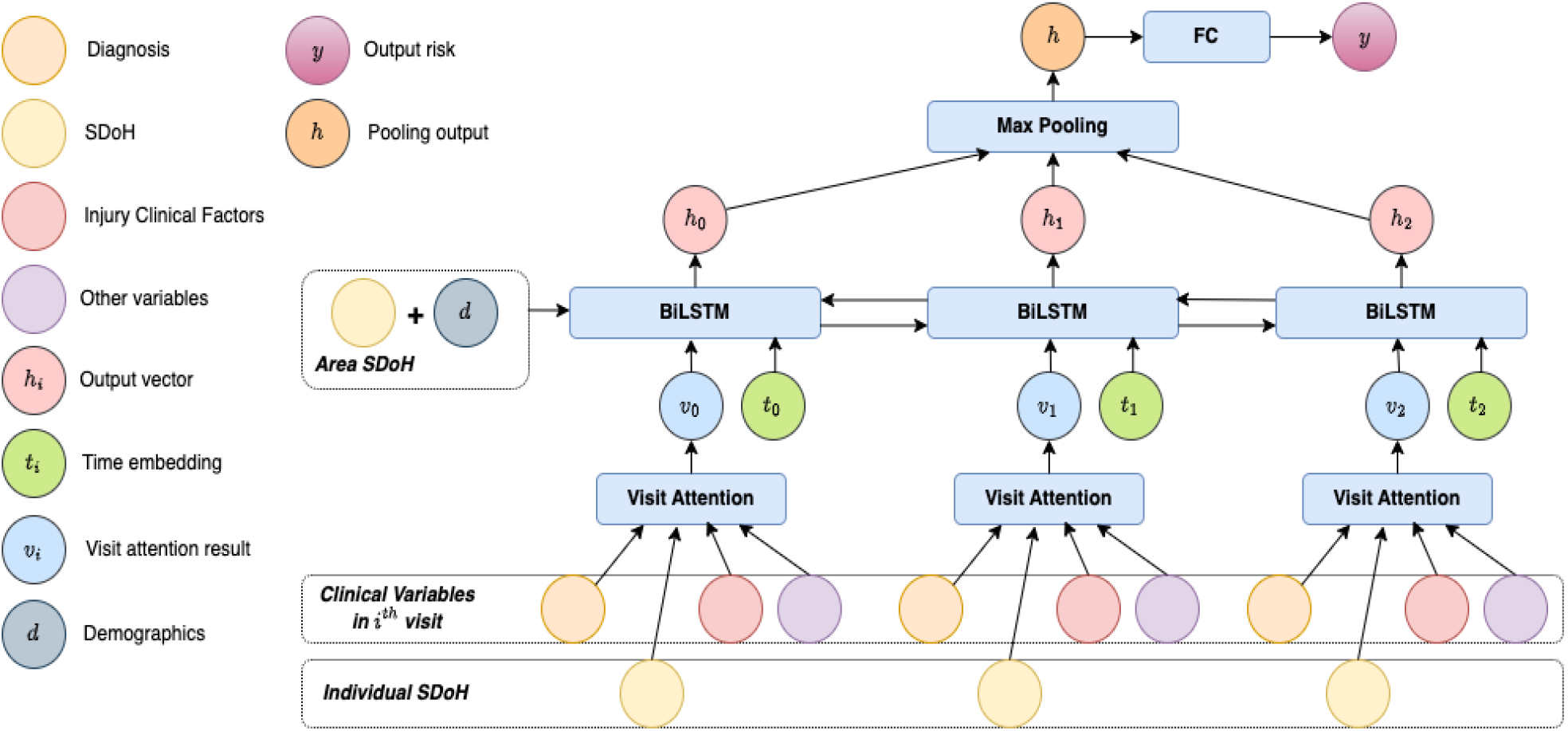
Illustration of our deep learning model based on a long short-term memory (LSTM) network.

### Model Training Settings

The dataset was randomly split into three subsets: a training set (60% of encounters), a validation set (20% of encounters), and a testing set (20% of encounters). The training and validation sets were used during model development, while the testing set was reserved for final performance evaluation. To prevent information leakage, the split was performed at the patient level across all encounters. Although this approach might cause slight discrepancies between the actual and theoretical number of encounters in each set, it ensured that data from the same patient did not appear in both the training and testing sets, thereby preserving the integrity and reliability of the model’s performance evaluation.

The models were trained for 40 epochs using the AdamW optimizer. To prevent overfitting, dropout and batch normalization were applied as regularization techniques. The training loop utilized the training set for learning, the validation set for model performance evaluation during training, and the testing set for final performance evaluation. The model with the lowest validation loss was saved, ensuring that the best model was retained for testing.

The learning rate for the AdamW optimizer was set to 1e-4, with weight decay set to 1e-2 for additional regularization. The Binary Cross-Entropy Loss (BCELoss) was used as the loss function for the models.

### Performance Metrics

Our binary classification task categorized patient encounters into either the mental health (MH) diagnosis positive class or the MH diagnosis negative class. We evaluated the models using the concepts of true positives (Tp), true negatives (Tn), false positives (Fp), and false negatives (Fn). For example, Tp represents the number of encounters correctly identified as having an MH diagnosis, while Fn represents the number of encounters with an MH diagnosis that were incorrectly classified as negative. We compared the models using various performance metrics: 1) Accuracy = (Tp+Tn)/(Tp+Tn+Fp+Fn); 2) Precision (P) = Tp/(Tp+Fp); 3) Sensitivity/Recall (R) = Tp/(Tp+Fn); 4) Specificity = Tn/(Tn+Fp); 5) F1-score = 2×P×R/(P+R); and 6) Area Under the Curve - Receiver Operating Characteristic (AUC-ROC).

## Results

### Details of the Dataset

Summary statistics of the patient demographics are presented in Table 2. Summary statistics of the dataset utilized in our study are presented in Table 3. There were 40,436 total encounters. The training set included 24,380 encounters. The validation set included 8140 encounters. These examples were utilized in experiments to fine tune hyper-parameters of different machine learning classifiers. The test set included 7916 encounters which were used for performance evaluation.

**Table 2.** Patient demographics.

|  | N | % |
| --- | --- | --- |
| Total | 15,259 | 100.0% |
| Age at concussion diagnosis (years) |  |  |
| Mean (SD) | 14.57 (1.87) |  |
| Min-Max | 11 - 17.99 |  |
| Gender |  |  |
| Female | 6,983 | 45.8% |
| Male | 8,276 | 54.2% |
| Race |  |  |
| Black | 2,447 | 16.0% |
| Asian | 200 | 1.3% |
| Multiple Race | 829 | 5.4% |
| Other/Unknown | 522 | 3.4% |
| White | 11,261 | 73.8% |
| Ethnicity |  |  |
| Hispanic or Latino | 453 | 3.0% |
| Not Hispanic or Latino | 14,540 | 95.3% |
| Other/Unknown | 266 | 1.7% |
| Insurance |  |  |
| Private | 7,140 | 46.8% |
| Medicaid | 4,513 | 29.6% |
| Self-pay | 335 | 2.2% |
| Unknown | 3,271 | 21.4% |
| Rural-Urban Location |  |  |
| Urban | 13,446 | 88.1% |
| Rural | 1,599 | 10.5% |
| Missing | 214 | 1.4% |

\*Patients aged 11-17 years with a confirmed concussion diagnosis between 1/1/2013 and 12/31/2022 (ten years).

**Table 3.** Statistics of the dataset used in the study.

|  | Total encounters | Encounters with MH diagnosis |
| --- | --- | --- |
| Overall | 40436 | 8761 |
| Training set (60%) | 24380 | 5273 |
| Validation set (20%) | 8140 | 1777 |
| Test set (20%) | 7916 | 1711 |

### Model Comparison

The performance of different models for predicting mental health diagnosis following the index concussion is presented in **Table 4**. The LSTM models generated the highest AUCs, accuracy, sensitivity, and F-1 scores. By adding SDoH variables to the data, the AUC of the LSTM model increased from 0.887 to 0.892, the accuracy increased from 0.873 to 0.883, the sensitivity increased from 0.523 to 0.580, and the F1-score increased from 0.641 to 0.683.

**Table 4.** Performance of the models on data with and without SDoH information.

| Performance metric (data without any SDoH) | LR | MLP | RNN | LSTM |
| --- | --- | --- | --- | --- |
| AUC-ROC | 0.837 | 0.865 | 0.867 | 0.887 |
| Accuracy | 0.852 | 0.863 | 0.867 | 0.873 |
| Precision | 0.728 | 0.88 | 0.887 | 0.829 |
| Sensitivity | 0.507 | 0.423 | 0.441 | 0.523 |
| Specificity | 0.947 | 0.984 | 0.985 | 0.97 |
| F1-score | 0.598 | 0.571 | 0.589 | 0.641 |
| Performance metric (data with both area-level and individual-level SDoH) | LR | MLP | RNN | LSTM |
| AUC-ROC | 0.833 | 0.861 | 0.882 | 0.892 |
| Accuracy | 0.854 | 0.872 | 0.876 | 0.883 |
| Precision | 0.734 | 0.881 | 0.897 | 0.83 |
| Sensitivity | 0.510 | 0.471 | 0.481 | 0.58 |
| Specificity | 0.949 | 0.983 | 0.985 | 0.966 |
| F1-score | 0.602 | 0.614 | 0.626 | 0.683 |
| LR: Logistic Regression; MLP: Multilayer Perceptron; RNN: Recurrent Neural Networks; LSTM: Long Short-Term Memory. |  |  |  |  |

### Interpretability of the LSTM Model

We computed the SHapley Additive exPlanations (SHAP)^45,46^ values to better understand the interpretability of our LSTM models. SHAP is a technique based on cooperative game theory and is widely used today to understand how different features impact the prediction of a model. **Figure 3** visualizes the importance and effect of different features on our LSTM model’s predictions using a SHAP summary plot. The x-axis represents features in the model that do not change over time, while the y-axis shows the SHAP values. A positive SHAP value indicates that a feature increases the prediction, whereas a negative SHAP value means it decreases the prediction. Each dot corresponds to a SHAP value for a specific feature in an individual prediction, reflecting how much that feature contributed to the prediction for a particular instance in the dataset. The spread of the dots along the y-axis demonstrates the range of SHAP values for each feature across all instances. For example, if many dots for a feature like “Age” have positive SHAP values, it suggests that increasing “Age” generally increases the model’s prediction. Conversely, if most SHAP values for a feature are negative, that feature tends to lower the model’s prediction. The variability in the spread of dots indicates how consistent or variable a feature’s impact is across different predictions; a wider spread suggests a more variable impact. Features such as “Urban/Rural” and “Race” exhibit a wide range of SHAP values, indicating their influence on predictions varies significantly between instances. On the other hand, features like “Pre-existing MH,” “Insurance,” and “Visit Lead Time” show significant SHAP values, underscoring their importance as predictors in the model.

**Figure 3.**
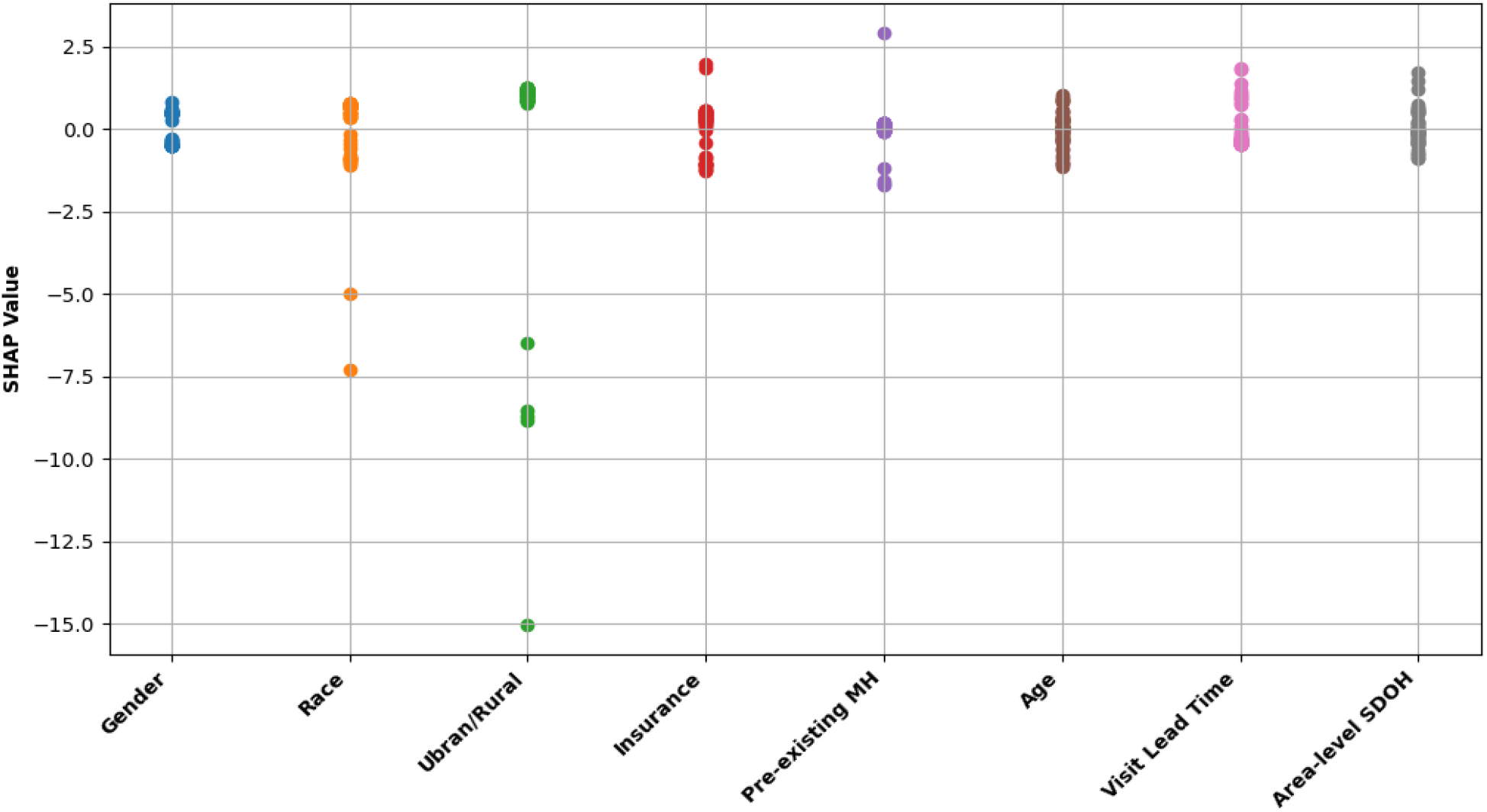
Illustration of Feature Contributions to BiLSTM Model Predictions Using SHAP Summary Plot. Visit Lead Time: number of days from index concussion diagnosis date to the first post-concussion visit date.

### At-the-visit Real-time Prediction

We selected an example patient to illustrate how our BiLSTM model makes predictions at each visit after the index concussion to aid in clinical decision making. **Figure 4** shows the visit trajectory of a male patient who lived in an area with low child opportunity index and had no historical mental health conditions. This patient had his first visit on day 1 and received a diagnosis of concussion. On day 2, he had his first post-concussion visit. At this visit, our LSTM model yielded a predicted risk of MH diagnosis as 0.431 (labeled as negative) while the ground truth was also negative (no MH diagnosis). On day 7, he had his second post-concussion visit. At this visit, additional clinical and injury variables that measure post-concussion symptoms were added to our model, yielding a predicted risk of MH diagnosis as 0.627 (labeled as positive) while the ground truth was also positive (a diagnosis of anxiety disorder was given). On day 14, he had his third post-concussion visit. At this visit, a new diagnosis of concussion was given, indicating he had a second concussion. Our BiLSTM model incorporated this new information along with additional post-concussion symptoms data, yielding a predicted risk of MH diagnosis as 0.839 (labeled as positive) while the ground truth was also positive. On day 23, he had his fourth post-concussion visit. At this visit, additional post-concussion symptoms data were added to our BiLSTM model, yielding a predicted risk of 0.855 (labeled as positive) while the ground truth was also positive.

**Figure 4.**
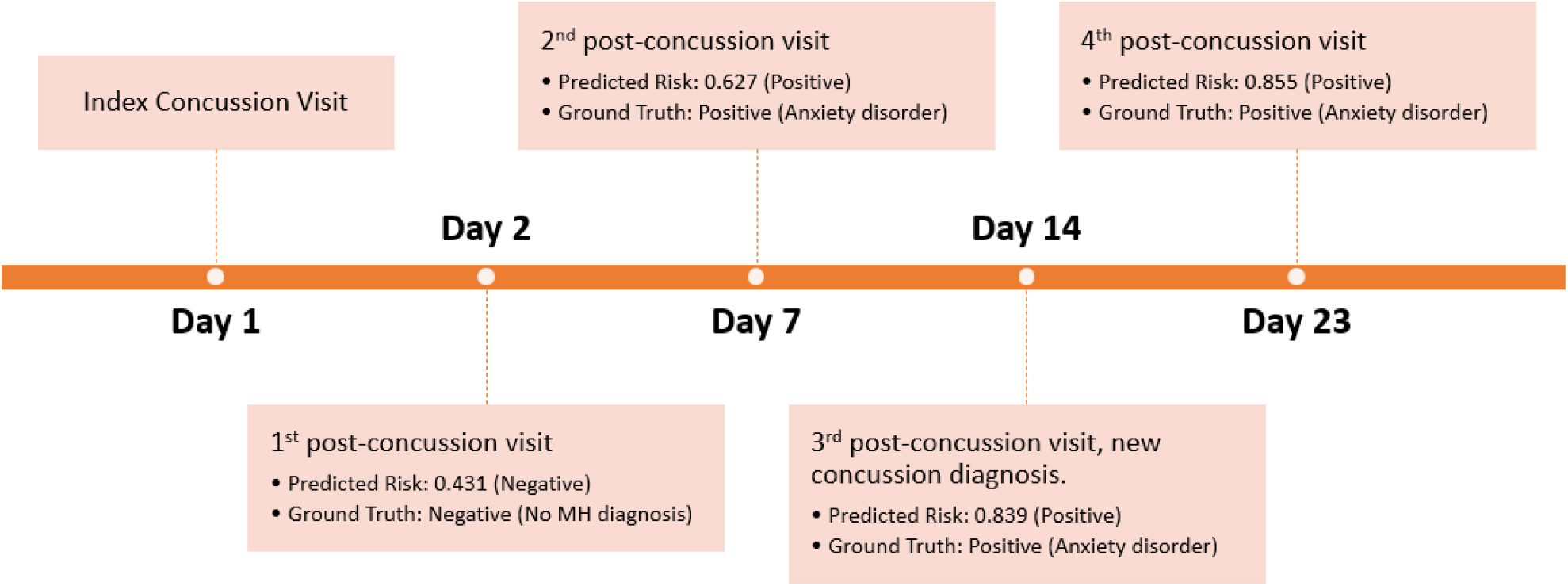
Visit trajectory of a male patient who lived in a metro area with low child opportunity index and had no historical mental health conditions.

## Discussion and Conclusions

The use of machine learning models for predicting outcomes in patients with TBI has garnered significant attention in recent years. In 2022, Pease et al. demonstrated that a deep learning model incorporating head CT scans and clinical information can effectively predict 6-month outcomes following severe TBI.^26^ Similarly, in 2018, Rau et al. found that artificial neural networks provided the most accurate predictions of mortality in patients with isolated moderate and severe TBI.^28^ In 2019, Feng et al. compared twenty-two machine learning models against logistic regression for mortality prediction in severe TBI patients in a single-center study, discovering that support vector machines outperformed logistic regression.^30^ However, these models primarily focus on more severe TBIs, often involve small sample sizes, and are typically applied to adult patient populations.

Our study is the first to develop an interpretable deep learning model using a bidirectional long short-term memory (BiLSTM) network to predict mental health sequelae following a concussion, a form of mild TBI, specifically in youth. When compared to other machine learning models, including Logistic Regression (LR), Multilayer Perceptron (MLP), and Recurrent Neural Networks (RNN), our BiLSTM model has demonstrated superior performance, achieving the highest AUC-ROC (0.887), accuracy (0.873), sensitivity (0.523), and F1 score (0.641). Incorporating social determinants of health (SDOH) data has further improved the BiLSTM model’s performance, resulting in an even higher AUC-ROC (0.892), accuracy (0.883), sensitivity (0.58), and F1 score (0.683). To enhance interpretability, we utilized SHAP values, making the model’s predictions more transparent. For instance, features like “Pre-existing mental health diagnoses,” “Insurance,” and “Visit Lead Time” were identified as significant predictors due to their high SHAP values. Conversely, features such as “Urban/Rural” and “Race” displayed a wide range of SHAP values, indicating their varying influence across different predictions. Moreover, unlike previous approaches that provide a one-time prediction of outcomes in patients with TBI, our model delivers predictions at each visit within 12 months following the index concussion. This real-time prediction capability allows clinicians to make immediate, visit-specific referrals for further treatment and interventions.

Our approach holds considerable promise for aiding in the early detection of mental health sequelae following concussion in youth at concussion clinics. While the dataset used in our study is from a single institution, we expect that our approach to be adapted by other institutions nationwide. Future research is necessary to enhance our training models with additional data and to evaluate our prediction models in real-world implementation studies.

## Data Availability

All data produced in the present study are available upon reasonable request to the authors

## Acknowledgements

This project was supported by National Institute of Mental Health funded Center for Accelerating Suicide Prevention in Real-world Settings (ASPIRES) Pilot Grant Program (P50MH127476). The content is solely the responsibility of the authors and does not necessarily represent the official views of the funder.

